# Alterations in aperiodic and periodic EEG activity in young children with Down syndrome

**DOI:** 10.1101/2024.05.01.24306729

**Authors:** McKena Geiger, Sophie R. Hurewitz, Katherine Pawlowski, Nicole T. Baumer, Carol L. Wilkinson

## Abstract

Down syndrome is the most common cause of intellectual disability, yet little is known about the neurobiological pathways leading to cognitive impairments. Electroencephalographic (EEG) measures are commonly used to study neurodevelopmental disorders, but few studies have focused on young children with DS. Here we assess resting state EEG data collected from toddlers/preschoolers with DS (n=29, age 13-48 months old) and compare their aperiodic and periodic EEG features with both age-matched (n=29) and cognitive-matched (n=58) comparison groups. DS participants exhibited significantly reduced aperiodic slope, increased periodic theta power, and decreased alpha peak amplitude. A majority of DS participants displayed a prominent peak in the theta range, whereas a theta peak was not present in age-matched participants. Overall, similar findings were also observed when comparing DS and cognitive-matched groups, suggesting that EEG differences are not explained by delayed cognitive ability.

## Introduction

Down syndrome (DS), caused by an extra full or partial copy of chromosome 21, is the most common genetic cause of intellectual disability, with most individuals exhibiting mild-moderate cognitive impairment^1^. In addition to a myriad of co-occurring medical conditions, children with DS often have significant language impairments and articulation challenges, motor delays with hypotonia, as well as learning and memory problems, with varying degrees of severity^2^. These cognitive and communication impairments can greatly impact quality of life, health outcomes, and adult independence. However, our understanding of neural mechanisms underlying cognitive profiles in DS is limited, preventing the development of effective therapies.

Electroencephalography (EEG) provides a non-invasive measure of network level brain activity and is well-tolerated in both children and adults with neurodevelopmental and genetic disorders^3^. Across early development, resting state EEG activity, as reflected in the EEG power spectrum, undergoes significant change^4–8^ as inhibitory networks and thalamocortical circuits are established and mature^9–12^.

Characterizing the resting state EEG power spectra in children with DS could provide insight into possible alterations in excitatory and inhibitory imbalance, as well as differences in brain oscillations that may impact sensory and cognitive processing.

Research in animal models of DS suggest altered function of inhibitory neurotransmitters, resulting in over-inhibition^13–17^. In contrast, human pediatric studies in DS have observed reduced GABA concentrations, suggesting there may be increased excitation at least early in development^18^. Accumulating evidence suggests that the slope of the EEG power spectra (also referred to as the aperiodic slope) reflects the excitatory-inhibitory (E/I) balance of the network, with a steeper slope associated with increased inhibition over excitation^19,20^. Thus, evaluating the aperiodic slope in children with DS could provide further evidence of altered E/I balance.

Oscillatory activity occurring in narrow frequency bands result from the coordination of inhibitory and thalamocortical network responses to sensory stimuli^21–25^, and thus, alterations in the developmental maturation of these systems could result in altered oscillatory activity. In DS, alterations in oscillatory activity have been observed in theta, alpha, and beta frequency bands (e.g.^26–29^). In adults with DS, multiple studies have observed increased power in slow frequencies (2-8Hz, delta and theta) and reduced alpha activity compared to neurotypical individuals^27,29,30^. Observations however have been mixed in higher frequencies^27,29–31^. Notably, very few studies have investigated EEG patterns in children with DS. Kaneko et al. 1996^32^ collected resting state EEG data in children aged 4-15 years with DS. As observed in adults, the pediatric DS group demonstrated increased delta and theta power, as well as decreased alpha power. No differences in higher frequencies were observed. Authors also observed that peak theta and peak alpha frequencies were lower in participants with DS compared to controls. Similarly, Katada et al. observed children with DS often presented with a dominant theta, rather than alpha peak, sometimes persisting into adulthood^28^. Even fewer studies have evaluated EEG in infants and toddlers with DS^26^ and therefore it is unclear whether similar differences in resting state activity are present early in development. Importantly, during early typical development as networks mature the peak frequency and amplitude of oscillatory activity changes; for example the dominant alpha rhythm observed in adulthood (∼10hz) initially occurs in the theta/low alpha range (∼5Hz) in infancy, and then gradually increasing in amplitude and peak frequency with age^5,6^. These oscillations are associated with cognitive processing^33^; for example, the strength of alpha oscillations is modulated by attention^34,35^, whereas oscillations in the beta range have been associated with sensory motor processing and high-order cognitive tasks^36,37^. Assessing differences in the development of oscillatory activity in young children with DS can provide insight into underlying neural mechanisms and altered network development.

To evaluate the slope of the power spectra and oscillatory activity, new methods have been developed to parametrize the spectra into aperiodic and periodic components^38^. The aperiodic component follows the 1/f power law distribution, defining the slope of the power spectrum. The periodic component is defined by the portions of the power spectrum rising above the aperiodic component, with peaks reflecting oscillatory activity in narrow frequency bands^33^. Growing evidence suggests that parameterization of the power spectra is important for accurate measurement of peak amplitude and frequency, and for interpretation of findings^39^. For example, increases observed in theta power in DS could be due to both increases in aperiodic activity and specific increases in periodic theta activity. Characterizing differences in the DS power spectra as it relates to both aperiodic and periodic activity is thus critical in understanding the underlying neurobiology of the disorder.

In this study, we aimed to characterize resting-state (non-task) EEG activity in a group of toddler-preschool aged children with DS (n = 29), as compared to either age-matched (n=29) or cognitive-matched (n=58) comparison groups. We hypothesized that alterations in both aperiodic and periodic components would be observed including increased aperiodic slope, increased periodic theta activity, and decreased periodic alpha activity.

## Methods

### Participants

This analysis includes infants and children with Down syndrome (mean age 29.0 ± 9.2 months, range 13-48 months) recruited as part of two separate studies collected in the same lab from 2019-2022. (IRB-P00018377, IRB-P00025806). DS participants all had Trisomy 21, a minimum gestational age of 30 weeks, a birth weight of >2000 gms, and their families spoke primarily English at home (>50%). Children with known neurological disorders (e.g. intraventricular hemorrhage, seizure disorder) were also excluded. DS participants were not excluded for diagnoses of autism (n=1), minimal-moderate hearing loss (n=6), visual impairment (n=9), or other common medical comorbidities occurring in DS such as congenital heart disease, asthma, and obstructive sleep apnea. Of the 34 enrolled participants with cognitive testing and EEG data collected at the same time point, 5 EEG files were excluded due to data quality issues (see EEG pre-processing and rejection criteria below). Therefore, a total of 29 participants were available for analysis across the two studies.

Both age-matched and cognitive-matched comparison groups were identified from co-occurring studies (IRB-P00018377, IRB P00025493) in the same laboratory. A cognitive-match comparison group allows us to assess whether EEG differences present between DS and age-matched groups represent delays in brain maturation. In this case we would expect that EEG features observed in DS are more similar to controls of similar developmental ability rather than similar age. Comparison group participants had a minimum gestational age of 35 weeks, a birth weight of >2000 grams, no known genetic or neurological disorders, and their families spoke primarily English at home (>50%). All age matches were within +/- 3 months for participants younger than 30 months (avg. 1.3 months, standard deviation (SD) =1.3 months) and +/- 6 months for participants 30 months or older (avg. 3.8 months, SD=1.8 months).

Cognitive matches were identified using scores on the Mullen Scales of Early Learning (MSEL^40^), a standardized developmental measure for children 0-69 months of age that was administered across all studies. Cognitive matches were identified using raw scores on nonverbal scales of the MSEL. First matches were identified using the Visual Reception scale, with two cognitive matches for each DS participant (avg. difference 0.6 points, SD=1.1 points). If multiple matches were available, we chose the match with the closer raw score on the Fine Motor subscale (avg. difference 1.4 points, SD=1.8 points). In order to identify age and cognitive matches for all DS participants, 11 TD children were used as both a cognitive match and an age match for different DS participants. Given this, age vs. cognitive matched comparisons were not statistically compared. Demographic data for all 3 groups are described in Table 1.

**Table 1.** Sample characteristics and EEG quality metrics.

|  | DS (n=29) | Age-Match (n=29) | Cog-Match (n=58) |
| --- | --- | --- | --- |
| Sex | 15M, 14F | 17M, 12F | 32M, 26F |
| Age (months) | 29.03 ± 9.19 | 29.79 ± 10.71 | 16.43 ± 5.45 |
| Ethnicity, n (%) |  |  |  |
| Not Hispanic or Latino | 29 (100) | 26 (90) | 55 (95) |
| Hispanic or Latino | 0 (0) | 2 (7) | 3 (5) |
| Not Answered | 0 (0) | 1 (3) | 0 (0) |
| Race, n (%) |  |  |  |
| White | 25 (86) | 21 (72) | 44 (76) |
| Black or African American | 1 (3) | 2 (7) | 1 (2) |
| Asian | 0 (0) | 1 (3) | 2 (3) |
| Mixed Race | 3 (10) | 4 (14) | 11 (19) |
| Not answered | 0 (0) | 1 (3) | 0 (0) |
| Household Income, n (%) |  |  |  |
| < \$30,000 | 2 (7) | 2 (7) | 1 (2) |
| \$30,000 - \$60,000 | 1 (3) | 1 (3) | 1 (2) |
| \$60,000 - \$100,000 | 8 (28) | 2 (7) | 6 (10) |
| More than 100,000 | 14 (48)<sup>a</sup> | 23 (79) | 48 (83) |
| Not answered | 4 (14) | 1 (3) | 2 (3) |
| Highest Level of Caregiver Education, n (%) |  |  |  |
| High school/GED | 3 (10) | 2 (7) | 1 (2) |
| Associate Degree | 3 (10) | 1 (3) | 0 (0) |
| Bachelor Degree | 8 (28) | 4 (14) | 11 (19) |
| Graduate/Professional degree | 15 (52) | 21 (72) | 46 (79) |
| Not answered | 0 (0) | 1 (3) | 0 (0) |
| EEG Quality Metrics |  |  |  |
| Number of Segments | 98.76 ± 30.65 | 109.28 ± 24.19 | 97.47 ± 37.64 |
| Percent Good Channels | 92.53 ± 4.70 | 92.81 ± 3.96 | 93.49 ± 3.39 |
| Percent ICs Rejected | 40.10 ± 11.33 | 30.99 ± 8.84 | 34.42 ± 10.26 |
| Mean Artifact Probability of Kept ICs | 0.14 ± 0.04 | 0.10 ± 0.04 | 0.12 ± 0.05 |
| Mullen Scales of Early Learning |  |  |  |
| Verbal Quotient | 57.37 ± 13.22 | 105.43 ± 21.42<sup>b</sup> | 99.16 ± 18.22 |
| Nonverbal Quotient | 60.50 ± 13.46 | 108.47 ± 14.33 | 106.62 ± 16.21 |
| Visual Reception Raw Score | 19.83 ± 6.41 | 33.28 ± 10.40 | 20.00 ± 6.22 |
| Fine Motor Raw Score | 18.72 ± 4.07 | 28.83 ± 7.96 | 18.93 ± 3.94 |

<sup>a</sup> 4 caregivers reported income >\$90,000 and were placed in the >\$100,000 category.
<sup>b</sup> N = 22, as one study providing age-matches did not complete the language sub-scales on the MSEL.

Note, studies spanned the COVID-19 pandemic and 10 participants in the DS group were seen after research operations resumed in 2021. All other participants were seen prior to the COVID-19 pandemic. Institutional review board approval was obtained prior to starting all studies. Researchers obtained written, informed consent from parents or guardians prior to their children’s participation.

### EEG Collection

For all participants across studies, resting-state, non-task related, EEG data was collected in a dimly lit, sound-attenuated, electrically shielded room. Participants sat in their seated caregiver’s lap or sat independently in a chair or high-chair, and caregivers were instructed by a research assistant to avoid social interactions or speaking with their child. Continuous EEG was collected while participants watched an abstract moving design on a computer monitor or if preferred for behavioral compliance, a silent video of their choosing on an iPad. EEG was recorded for between two and five minutes, depending on compliance, using a 128-channel Hydrocel Geodesic Sensor Net (Electrical Geodesics Inc., Eugene, OR) with electrooculographic electrodes removed, connected to a NetAmps 300 amplifier (Electrical Geodesics Inc.). Data was sampled at either 1000Hz or 500Hz depending on the study and referenced online to a single vertex electrode (Cz).

### EEG pre-processing

Raw Netstation (Electrical Geodesics, Inc) files were exported to MATLAB (MathWorks) for pre-processing and power analysis using the Batch EEG Automated Progressing Platform (BEAPP^41^) with integrated Harvard Automated Preprocessing Pipeline for EEG (HAPPE^42^). Each EEG underwent a 1Hz high-pass and 100Hz low-pass filter. Data was resampled to 250Hz and then run through the HAPPE module which included 60Hz line noise removal, bad channel rejection, and artifact removal using combined wavelet-enhanced independent component analysis (ICA) and Multiple Artifact Rejection Algorithm (MARA^43,44^). Given the short length of EEG recordings, 36 of the 128 channels were used for ICA/MARA: (Standard 10-20 electrodes: 22, 9, 33, 24, 11, 124, 122, 45, 36, 104, 108, 58, 52, 62, 92, 96, 70, 83; Additional electrodes: 28, 19, 4, 117, 13, 112, 41, 47, 37, 55, 87, 103, 98, 65, 67, 77, 90, 75). After artifact removal, channels removed during bad channel rejection were interpolated, data was then referenced to the average reference and detrended to the signal mean. Processed data was then segmented into 2-second segments, and HAPPE’s amplitude and joint probability criteria were used to reject segments with retained artifact.

### EEG rejection criteria

HAPPE quality measures were used to reject full EEG recordings from final analysis. EEGs were fewer than 20 segments (40 second of total EEG), percent good channels <80%, percent independent components rejected >80%, mean artifact probability of components kept was

<0.3, and percent variance retained <25%. Table 1 shows quality metrics across groups.

### EEG Power Spectra Analysis

The power spectral density (PSD) was calculated at each electrode, for each two second segment, using a multitaper spectral analysis^45^ using three orthogonal tapers. For each electrode, the PSD was then averaged across segments and then averaged across electrodes grouped into four regions of interest (frontal, temporal, central, and posterior – Supplemental Figure 1). The power spectra was then further parametrized using SpecParam v1.0.0^38^ (https://github.com/fooof-tools/fooof; in Python v3.6.8) in order to evaluate differences in aperiodic and periodic components of the power spectra. The SpecParam model was used in the fixed mode (no spectral knee) evaluating spectra between 2-55Hz, with *peak_width_limits* set to [0.5, 18.0], *max_n_peaks* = 7, and *peak_threshold* = 2). Mean R^2^ for the full sample was 0.997 (SD = 0.004; range 0.962-0.999).

The aperiodic 1/f signal can be described by its offset and slope, both provided by SpecParam. Here we define the aperiodic offset as the aperiodic power at 2.5Hz, as we have observed high levels of error in SpecParam estimates at frequencies below 2.5Hz^4^. To determine the periodic power spectra, the SpecParam estimated aperiodic signal was subtracted from the absolute power spectrum. To further characterize theta, alpha, and beta peaks, the periodic power spectra was smoothed using a savgoal filter (scipy.signal.savgol_filter, window length = 101, polyorder = 8), and then peak maxima were identified within the following frequency ranges: theta (4-6.5Hz), alpha (6.5-12Hz), low beta (12-20Hz), and high beta (20-30Hz) range. Periodic power was calculated using the integral of the periodic spectra for the following frequency ranges: theta (4-6Hz), low beta (12-20Hz), high beta (20-30Hz), and gamma (30-45Hz).

### Statistical Analyses

Group differences in the power spectra were examined using a non-parametric clustering method, controlling for multiple comparisons using Monte Carlo estimation with 10,000 permutations^46^, employed with MNE-Python^47^. Group differences in EEG measures were statistically compared using T-test, or Mann Whitney if data was not normal in distribution. Statistical assessment of group differences in frequency band power (Figure 3) were corrected for multiple comparisons (25 comparisons, 5 ROIs and 5 power variables) using the Bonferroni correction method.

## RESULTS

Absolute, aperiodic, and periodic power spectra for DS, age-matched, and cognitive-matched groups are shown for whole brain regions of interest (ROI), as well frontal, central, temporal, and posterior ROIs (**Figure 1**). The most prominent differences were observed in the periodic spectra, with differences identified across multiple frequency bands. We used a non-parametric clustering method, controlling for multi-comparisons to identify significant differences in the power spectra between (1) DS and age-matched, and (2) DS and cognitive-matched groups (**Table 2**). Below we further characterize group differences in aperiodic activity, and periodic features in theta, alpha, and beta frequency bands.

**Fig 1.**
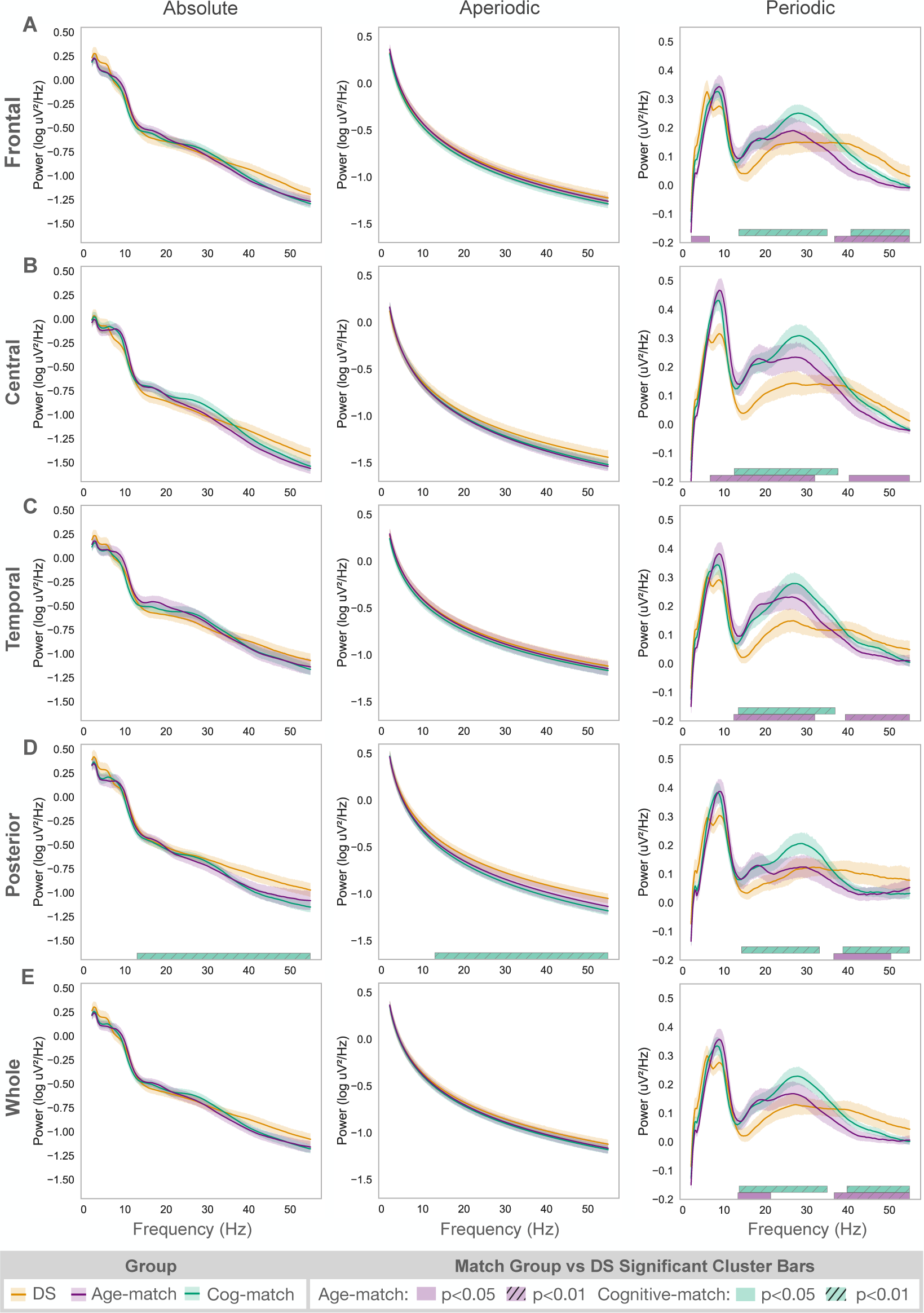
Absolute, aperiodic, and periodic activity for Down syndrome (yellow) and age-matched (purple) and cognitive-matched (green) comparison groups are shown for four regions of interest along with whole brain averages. Identified statistically significant clusters are shown as horizontal bars defined by the cluster’s frequency band. Shaded areas along spectra describe 95% confidence intervals.

**Table 2.** Periodic power spectra clusters with significant group differences.

| Region of Interest | DS vs. Age-Match |  | DS vs. Cog-Match |  |
| --- | --- | --- | --- | --- |
|  | Frequency range (Hz) | Cluster p-value | Frequency range (Hz) | Cluster p-value |
| Frontal | 2-6.5 | 0.03 | 13.5-35 | 0.001 |
| Frontal | 36.7-54.9 | 0.0005 | 40.6-54.9 | 0.006 |
| Central | 6.6-32 | 0.0006 | 12.4-37.6 | 0.0001 |
| Central | 40.2-54.9 | 0.01 | -- | -- |
| Temporal | 12.3-32 | 0.001 | 13.4-36.9 | 0.0001 |
| Temporal | 39.3- 54.9 | 0.009 | -- | -- |
| Posterior | 36.5-50.4 | 0.03 | 14.2-33.1 | 0.007 |
| Posterior | -- | -- | 38.7-54.9 | 0.008 |
| Whole | 13.3-21.3 | 0.04 | 13.6-35 | 0.001 |
| Whole | 36.6-54.9 | 0.001 | 39.7-54.9 | 0.006 |

### Aperiodic activity differences

Non-parametric cluster analysis identified a significant cluster between 13-55 Hz in posterior electrodes between DS and cognitive-matched groups. Further analysis of aperiodic slope identified significant differences between DS and both age- and cognitive-matched groups **(Figure 2A**). Specifically, the DS group exhibited lower slope compared to the age-matched group in frontal (t(56) = 2.08; p<0.05), central (t(56) = 3.56; p<0.001), and posterior (p<0.05) ROIs, and lower slope compared to the cognitive-matched group in central (t(85) = 3.66; p<0.001), and posterior (t(85) = 3.81; p < 0.001) ROIs. No differences between groups were observed for aperiodic offset.

**Fig 2.**
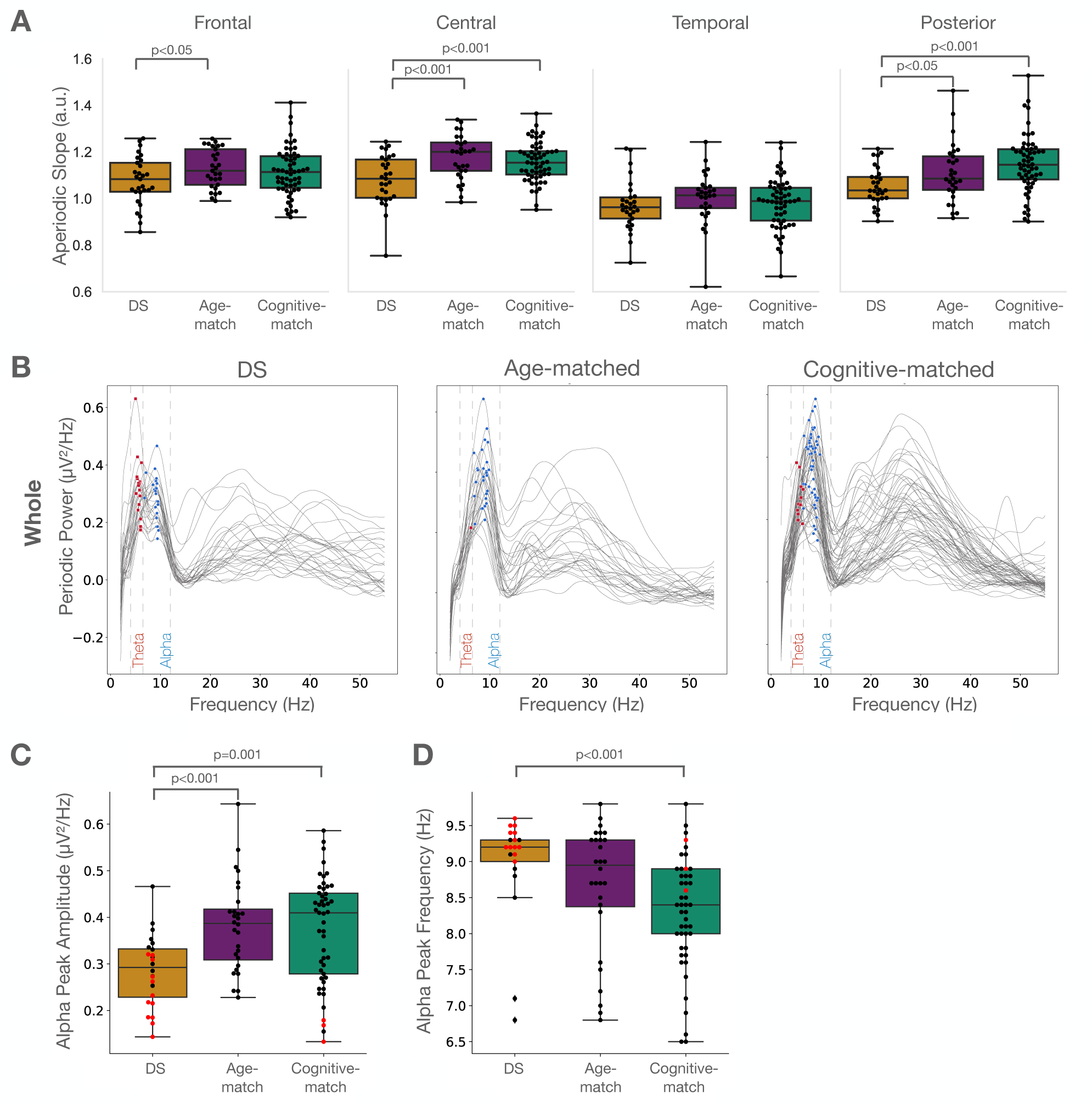
**(A)** Aperiodic slope between DS, age-matched, and cognitive-matched groups, across 4 regions of interest. **(B)** Individual periodic power spectra from electrodes across the whole brain. Identified peaks between 4-6.5Hz (theta) are shown in red, and peaks between 6.5-12Hz (alpha) are shown in blue. **(C-D)** Group differences in alpha peak amplitude and alpha peak frequency.

### Differences in periodic theta activity in DS group

Cluster analysis revealed a significant frontal cluster between 2-6.5Hz (p<0.05) for the DS vs. age-matched comparison. No other significant clusters were identified in the theta range. However, visual evaluation of both averaged and individual periodic spectra suggested two distinct peaks across the theta and alpha frequency bands (4-12Hz) unique to the DS group **(Figure 1, 2B).** Quantification of the number of peaks between 4-12Hz using whole brain ROI revealed that 41.4% (12/29) of the DS group had peaks in both the theta and alpha ranges. In contrast, no individual in the age-matched control group and 5.2% (3/58) of the cognitive-matched controls exhibited peaks in both the theta and alpha ranges. A chi-squared analysis revealed a significant difference between the number of individuals who exhibit a peak in both the alpha and theta ranges in a DS versus age-matched comparison (χ^2^ = 12.7; p<0.001) and DS verses cognitive-matched comparison (χ^2^ = 15.3; p<0.0001). Furthermore, 58.6% (17/29) of the DS group exhibited a theta peak (4-6Hz), while only 3.4% (1/29) of age-matched (χ^2^ = 18.1; p<0.0001), and 20.7% (12/58) of cognitive-matched groups (χ^2^ = 10.9; p<0.001) had a theta peak, suggesting some individuals in DS and cognitive-matched groups exhibited a peak in the theta but not in alpha band.

T-test revealed the whole brain theta periodic power for the DS group was significantly greater than the age-matched group (Figure 3A), but was not significantly different from the cognitive-matched group. ROI analysis showed greatest differences in theta power between the DS and age-matched groups in frontal regions (Figure 3A).

**Fig 3.**
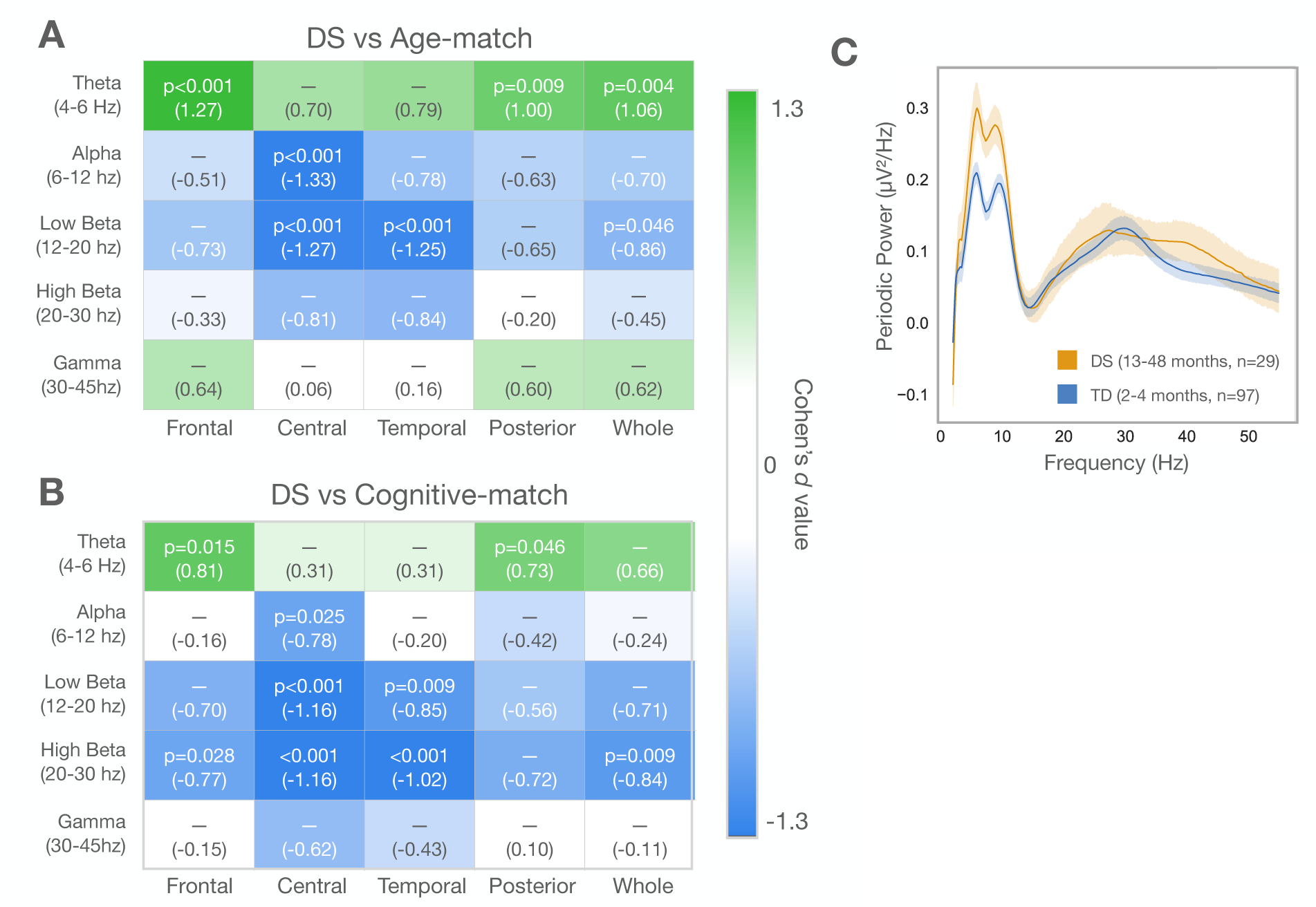
Heatmap of statistical Cohens’ d effect size of t-test comparison of difference in periodic power of five frequency bands between **(A)** DS and Age-matched groups and **(B)** DS and Cognitive-matched groups. p-value shown with effect size in parentheses. **(C)** Periodic power of DS group compared to a cohort of typically developing infants.

### Alpha periodic power differences

Cluster analyses identified a significant cluster between the DS and age-matched groups spanning the alpha band for central (Table 2), but not other regions of interest. We next assessed differences in whole brain alpha peak amplitude and frequency, defining alpha as between 6.5-12Hz (Figure 2C). As described above, not all individuals with DS had an alpha peak. The DS group showed significantly lower alpha peak amplitude (mean = 0.29 µV/Hz, n = 24) than the age-matches (mean = 0.38 µV/Hz, t(50) =3.87; p<0.001, n = 28) and cognitive-matches (mean = 0.37 µV/Hz, t(71) = 3.39; p=0.001, n = 49). As expected, given differences in age between groups, alpha peak frequency was significantly higher in the DS group (mean = 9.0Hz) compared to the younger in age cognitive-match group (mean = 8.3Hz; t(71) = -3.67; p<0.001) while there was no significant difference between DS and age-match groups (mean = 8.7Hz; t(50) = -1.55; p=0.13). The majority of individuals in the DS group with an alpha peak had a peak between 8.5-9.75 Hz (Figure 2D). When further restricting the DS group to those with two distinct peaks across the theta-alpha range, the alpha peak frequency range narrows to 9-9.75 Hz (Figure 2C,D, red). When comparing periodic alpha power between groups, significant differences were only observed for the central ROI (Figure 3A, B)

### Beta and Gamma periodic power differences

Cluster analyses identified significant differences between DS and cognitive-matched periodic spectra in the beta range (∼13-35Hz, Table 2, Figure 1) across all ROIs, with the DS group showing lower power than the cognitive-matched group. The DS group also showed reduced periodic power in the low beta range compared to the age-matched group specifically for central and temporal ROI’s (Figure 3A). Assessment of peak differences, showed that only six DS participants (20.7%) exhibited a low beta peak, while 62.1% (18/29) of age-matched participants (χ^2^ = 8.6; p<0.005), and 48.3% (28/58) of cognitive matched participants (χ^2^ = 5.1; p<0.05) showed a low beta peak. Differences were also observed between DS and cognitive-matched groups. Low beta (12-20Hz) power was significantly lower in central and temporal ROIs compared to both control groups (Figure 3A,B). The DS group also exhibited significantly lower high beta compared to cognitive-matched participants across frontal, central, and temporal electrodes (Figure 3B).

Cluster analyses also identified significant clusters in the gamma range (35-55Hz), with the DS group showing higher whole brain gamma power compared to both age-matched and cognitive-matched groups (Table 2). T-test group comparison of gamma power (30-55Hz) however did not show any significant differences after correcting for multiple comparison. This difference in statistical findings may be the result of how the beta and gamma frequency bands are defined and group differences in the “shape” of the periodic power spectra, with the DS group have a flattened and broader beta peak that extends in the gamma range.

## DISCUSSION

In this study we compared resting state EEG power spectra in young children with DS compared to both age- and cognitive-matched comparison groups. Instead of traditional assessments of absolute or relative power, we separately analyzed aperiodic activity (reflecting broadband background signal) and periodic activity (reflecting narrowband oscillatory components) allowing for a more accurate characterization of group differences in the power spectra^39,48^. Using this methodology, we observe four major differences between DS and age-matched comparison groups. First, aperiodic slope was significantly lower in children with DS, suggesting an increased E/I ratio. Second, the majority of children with DS displayed a peak in the theta range, whereas no theta peak was present in any age-matched participants, leading to significantly higher periodic theta power for the DS group. Third, alpha power and alpha peak amplitude were both reduced in the DS group. Fourth, significantly fewer DS participants had a low beta peak compared to age-matched groups. Notably, these differences were also observed when comparing with cognitive-matched comparison groups, suggesting that differences are specific to the genetic disorder rather than cognitive ability. Below we discuss how the above findings relate to prior research in DS and typical development.

### Aperiodic activity in DS

In our sample, children with DS exhibited a lower aperiodic slope compared to both cognitive- and age-matched control groups. While statistically significant differences between DS and age-matched control groups were not observed across whole brain electrodes, significant differences were observed for frontal, central, and posterior electrodes. There is growing evidence that aperiodic slope is an indirect measure of E/I balance, with lower slope (flatter aperiodic curve) indicative of a higher E/I ratio^19,49^. Our findings suggest there is increased excitation over inhibition in toddler/preschool aged children with DS compared to cognitively- and age-matched children. These findings contrast with research in mouse models of DS, where excessive inhibitory GABA-mediated signaling has been observed, and treatments using GABA antagonists have improved cognitive deficits in mice^13,50^. However, others have found that GABA-mediated signaling may actually be excitatory in DS^51^, and that in the fetal frontal cortex^18^ and temporal lobes^52^ of children with DS, GABA levels are reduced. Altered GABAergic signaling has been implicated in a number of neurodevelopmental disorders^53^; thus, future research investigating aperiodic slope in both animal models and humans across development may help tease apart the role of GABAergic signaling and E/I balance in DS.

### Periodic theta and alpha findings

Both theta and alpha oscillations are thought to be regulated by thalamocortical circuitry^34,35^, and across early development multiple studies have observed a shift in the dominant oscillatory frequency from a ∼5Hz theta range at five months of age, to a ∼8Hz alpha peak at two years, to a “mature” 10Hz alpha peak by adolescence^5,6,8,54^. This shift in peak frequency with age is also accompanied by an increase in alpha peak amplitude. In DS, both increases in theta activity and decreases in alpha activity have been consistently reported, albeit mostly in studies of adults (reviewed in ^26^). Our findings in young children with DS provide additional developmental context to these observations.

Here, we observe that more than half of children with DS in our sample exhibited a theta peak, leading to an increase in theta power. However, this finding cannot simply be interpreted as a delayed shift in peak frequency, as over 40% of the DS group displayed *both* theta and alpha peaks, with the majority of alpha peak frequencies falling between 9-10Hz. In one of the few longitudinal studies of EEG in DS, Katada et al.^28^ observed that at the youngest ages in their sample (four years old), the dominant peak frequency of the absolute power spectra was in the theta range, and this was most robust in frontal and central electrodes. Katada et al. also reported the presence of both theta and alpha peaks into adulthood. Interestingly the 4-5Hz activity was so elevated in young adults, authors excluded further analysis of this frequency range as they felt it could reflect artifact. Our similar findings in children suggest that a high theta peak amplitude and presence of both theta and alpha peaks reflects altered neurodevelopment in DS, rather than artifact. Several aspects of theta peaks observed in preschoolers with DS are remarkably similar to what we have previously reported in very young neurotypical infants. In a longitudinal analysis of developmental trajectories of periodic power we have previously observed the presence of both theta and alpha peaks in ∼70% of infants between 2 and 4-months-old, however by 6-months-old, the majority of infants displayed a single peak, usually in the theta range that then increased in frequency with age^4^. In Figure 3C, we show the similarities between average periodic power from the DS group and average periodic power from 2-4 month-olds. Together, these findings suggest that persistent increases in theta activity in individuals with DS may represent the sustained presence of a normally transient step in neurodevelopment.

Like previous studies in DS children and adults, we also observed a decrease in alpha activity as measured by peak alpha amplitude. The mature alpha rhythm observed in adults is thought to support cognitive functions such as attention and memory^34,35^, and in DS adults, reduced alpha activity has been associated with greater cognitive deficits^29^. However, the atypical presence of both theta and alpha peaks in our pediatric DS group suggests that the developmental maturation of the alpha peak in DS may be abnormal. Indeed, the expected positive association between alpha peak frequency and age are only observed in the age-matched comparison group and not the DS group (Supplemental Figure 2). The alpha rhythm is modulated both by thalamocortical activity and cholinergic signaling^55^. It has therefore been hypothesized that reductions in alpha amplitude observed in individuals with DS could reflect both alterations in thalamocortical development^56^, as well as altered cholinergic pathways which have been observed in older adults with DS and is implicated in Alzheimer’s disease^57^. Further research in larger samples of children is needed to investigate how alterations in both theta and alpha activity are associated with cognition in DS.

### Periodic beta and gamma findings

Compared to the age-matched comparison group, we observed reduced activity across the periodic beta range, and increased activity in the gamma range. Reductions in the beta band have also been observed in individuals with DS^26,29,56^ although findings have been mixed. Differences between previous studies could be due to age dependent changes in beta activity, as well as effects of medication (e.g.

GABAergic medications) on beta activity. In our sample, only six participants with DS exhibited a low beta peak, which was markedly lower than both age-matched and cognitive-matched groups. Prior longitudinal analysis of neurotypical infants suggests that by age 18 months, 50% of infants have a low beta peak^4^. Future research across a broader age range of individuals with DS will be critical in understanding the developmental origin and persistence of observed differences across development.

Significant differences were also observed between DS and the younger cognitive-matched groups in the high beta band, with the cognitive-matched group have higher beta activity. This is partially due to expected age differences between the two groups (DS mean age = 29 months, Cog-match mean age = 16 months), as neurotypical children between 6 and 20 months exhibit strong high beta activity with peak frequency between 26-30Hz, which then decreases in amplitude and peak frequency by 36 months^4^. The presence of differences between DS and younger cognitive-matched groups suggest that alterations in DS are not simply due delayed brain maturation, but instead reflect altered brain development.

Increases in gamma activity in the DS compared to the age-matched group were also observed. Few studies have previously investigated gamma activity in DS. Cortical gamma oscillations are generated by parvalbumin (PV) inhibitory neurons and therefore alterations observed in DS could indirectly reflect differences in excitatory and inhibitory (E/I) balance^58^. Indeed, elevated gamma activity has been observed in other neurodevelopmental disorders hypothesized to have altered E/I balance, including Fragile X Syndrome (FXS) and autism^59–62^.

### Limitations and Future Directions

The sample size, while relatively large compared to other studies in DS, is still small. However, as seen in individual plots in Figure 2B, our observations are consistent on the individual level. Given our sample size, the current analysis does not include any assessment of associations between EEG measures and cognitive, language, or behavioral measures. While differences were observed between groups, whether these differences are associated with cognitive deficits observed in children with DS is still unknown. Future research, with larger samples, is required to adequately assess brain-behavior associations.

## Summary

In summary, resting-state EEG measures from this study in young children with DS identified alterations in aperiodic slope, elevated theta activity with persistent presence of a theta peak, as well as reduced alpha activity. Future studies examining developmental trajectories of aperiodic and periodic activity starting in infancy will provide additional insight into the developmental neural mechanisms underlying altered activity.

## Data availability

Consents obtained from human participants prohibit sharing of de-identified individual data without data use agreement in place. Please contact the corresponding author with data requests.

## Acknowledgements

We thank all the children and families who generously participated in this research. We thank all the research staff involved in participant recruitment, data collection, and database administration.

## Author Contributions

**Carol Wilkinson:** Conceptualization, Project administration, Supervision, Methodology, Software, Formal analysis, Data Curation, Writing – Original Draft, Visualization. **McKena Geiger:** Methodology, Formal analysis, Writing – Original Draft, Visualization. **Sophie Hurewitz:** Methodology, Formal analysis, Writing – Original Draft, Visualization. **Katherine Pawlowski**: Supervision, Writing – Review & Editing. **Nicole Baumer:** Resources, Writing – Review & Editing, Supervision, Project administration, Funding acquisition.

---

**Supplemental Figure 1.**
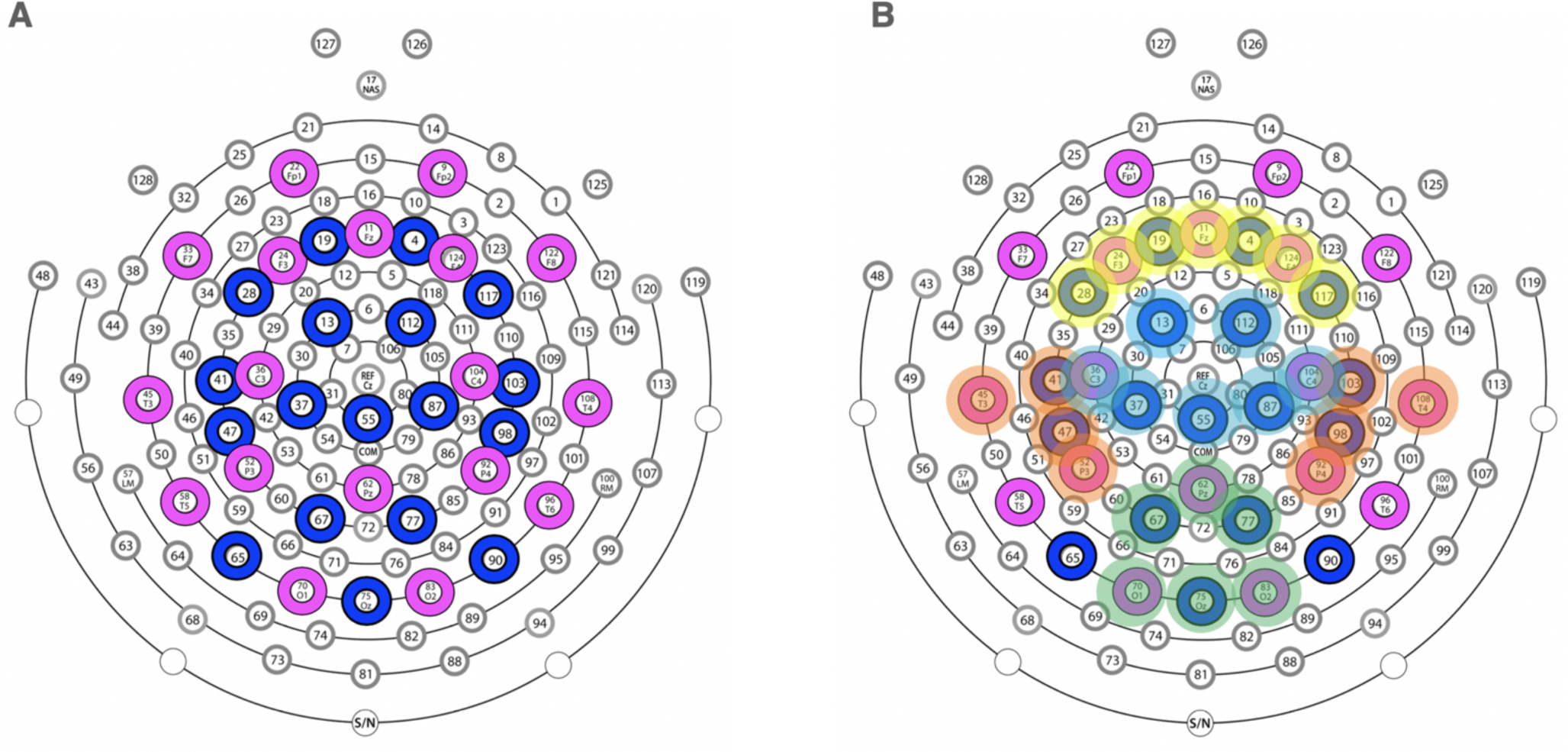
Electrode layout: (A) 128-channel Hydrocel Geodesic Sensor Net. Pink circles denote 10-20 electrodes, and blue circles denote the additional electrodes included in ICA and MARA steps of pre-processing. (B) Electrodes averaged for frontal (yellow), central (blue), temporal (orange), and posterior (green) regions of interest.

**Supplemental Figure 2.**
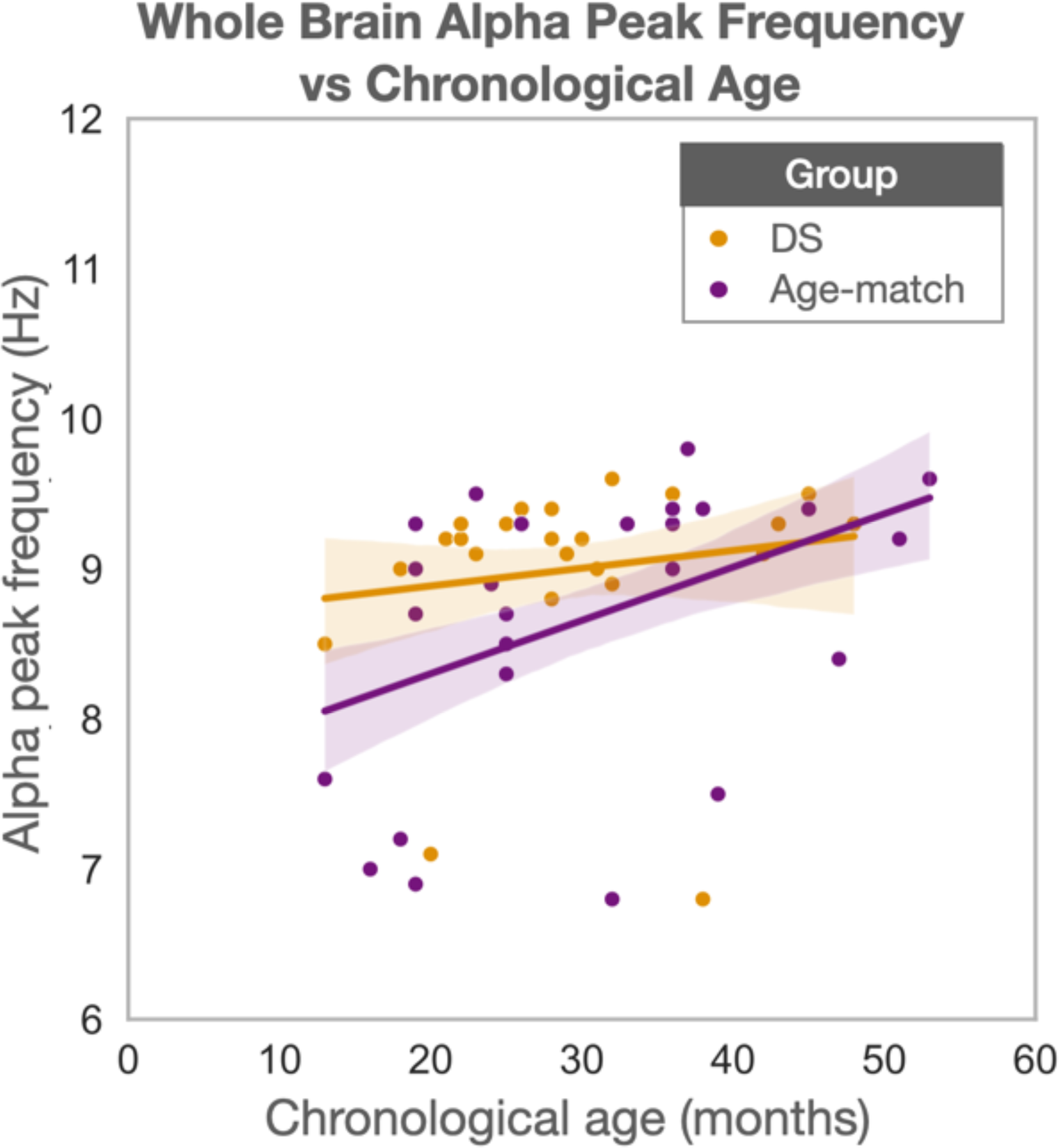
Whole brain alpha peak frequency vs chronological age for DS and age-match groups. : scatter plot of participant age verses alpha peak frequency for DS (orange) and age-match (purple) participants with regression lines. Pearson correlation coefficient revealed no significant correlation between chronological age and alpha peak frequency in the DS group, but showed a signification correlation within the age-matched group (p= 0.02, *r=* 0.44).

